# Improving Interprofessional Collaboration: Building Confidence Using A Novel HIV Curriculum For Healthcare Workers Across Sub-Saharan Africa

**DOI:** 10.1101/2023.11.14.23298522

**Authors:** Judy Khanyola, Mike Reid, Rand Dadasovich, Miliard Derbew, Ian Couper, Edward T. Dassah, Maeve Forster, Onesmus Gachuno, Clara Haruzivishe, Abigail Kazembe, Shayanne Martin, Mmoloki Moltwantwa, Keneilwe Motlhatlhedi, Kien Alfred Mteta, Nisha Nadesan-Reddy, Fatima Suleman, Catherine Ngoma, Georgina N. Odaibo, Roy Mubuuke, Deborah von Zinkernagel, Elsie Kiguli-Malwadde, David Sears

**Author notes:** Corresponding author Michael Reid, University of California, San Francisco, Institute for Global Health Sciences, 550 16th Street, Third Floor, San Francisco, CA 94158, USA. Contributed equally (Judy Khanyola, Mike Reid). Contributed equally (Elsie Kiguli-Malwadde, David Sears).

## Abstract

The 21st century presents significant global health challenges that necessitate an integrated health workforce capable of delivering person-centered and integrated healthcare services. Interprofessional collaboration (IPC) plays a vital role in achieving integration and training an IPC-capable workforce in Sub-Saharan Africa (SSA) has become imperative. This study aimed to assess changes in IPC confidence among learners participating in a team-based, case-based HIV training program across diverse settings in SSA. Additionally, it sought to examine the impact of different course formats (in-person, synchronous virtual, or blended learning) on IPC confidence. Data from 20 institutions across 18 SSA countries were collected between May 1, 2021, and December 31, 2021. Logistic regression analysis was conducted to estimate associations between variables of interest and the gain in IPC confidence. The analysis included 3,842 learners; nurses comprised 37.9% (n=1,172) and physicians 26.7% (n=825). The majority of learners (67.2%, n=2,072) were pre-service learners, while 13.0% (n=401) had graduated within the past year. Factors significantly associated with increased IPC confidence included female gender, physician cadre, completion of graduate training over 12 months ago, and participation in virtual or in-person synchronous workshops (p<0.05). The insights gained from this analysis can inform future curriculum development to strengthen interprofessional healthcare delivery across SSA.

## Introduction

Despite substantial research indicating that interprofessional education (IPE) enables effective collaborative practice leading to improved health outcomes and stronger health systems,[1-6] there is a paucity of data describing strategies to enhance interprofessional education and collaborative practice (IPECP) in Sub-Saharan Africa (SSA).[7] In this paper, we evaluate the impact of a multi-country HIV IPE training program to improve learner confidence in interprofessional collaboration (IPC). The program, which included both in-service and pre-service learners from 18 SSA countries, assessed the impact of three different formats for interprofessional learning on learner IPC confidence: in-person workshops, synchronous virtual workshops, and online workshops that blended asynchronous and synchronous components. By examining changes in IPC confidence after completion of the training and determining if course format had any impact on IPC confidence change, this analysis sought to inform curriculum development to strengthen interprofessional collaborative practice across SSA.

## Background

By 2030, it is estimated that SSA will face a shortage of 6 million healthcare workers, creating an urgent need to train a health workforce capable of responding to the numerous health challenges experienced by the region.[8] To address this need, diverse team-based interprofessional education (IPE) programs can play a critical role, preparing health professions trainees to deliver high-quality care in Africa, particularly in areas with a high burden of HIV.[9-12] One such educational initiative is the STRengthening InterProfessional Education for HIV (STRIPE HIV) program,[9] which aims to optimize team-based HIV care through the delivery of a curriculum consisting of case-based lessons targeting pre-service learners and early career in-service health professionals in high HIV burden countries across SSA. The program leverages a continent-wide network of health professions training institutions and affiliated health educators that are part of the African Forum for Research and Education in Health(‘AFREHealth’).[13]

## Methods

### Study Design and Subjects

The study was conducted using data from the STRIPE HIV program, which aimed to optimize team-based HIV care through a curriculum consisting of 17 case-based modules targeted at pre-service learners and early career in-service health professionals in high HIV-burden countries across Sub-Saharan Africa. The program included learners from 20 institutions in 18 countries. The curriculum was delivered in three formats: in-person workshops, virtual workshops (conducted synchronously on Zoom), or a blended online course (which included an online asynchronous component followed by a virtual synchronous component). The choice of learning format, which of the 17 case-based modules are taught, and the mix of learners engaged in the training was determined by local collaborating institutions and has been described previously. [9, 10]

### IPC Confidence Assessment

All learners who completed both the pre-test and post-test assessment for any of the 17 modules between May 1, 2021, and December 31, 2021, were included in the study, regardless of professional cadre (physician, nurse or midwife, pharmacist, laboratorian, etc.), stage of professional development (pre-service, less than 12 months of graduation, or greater than 12 months post-graduation) or geographic setting. These assessments captured learners self-reported confidence in IPC on a 4-point Likert scale prior to the course and following course completion, with one confidence assessment question for each module completed. The primary objective of this analysis was to examine changes in IPC confidence after completion of the curriculum and to determine if course format had any impact on IPC confidence change.

### Statistical Analysis

Descriptive statistics were used to summarize the demographic characteristics of learners and mean changes in IPC confidence for each demographic and each module. Individual learners were classified as having a gain in IPC confidence if their post-course self-reported IPC confidence score was higher than their pre-course self-reported IPC confidence score. We estimated associations between variables of interest and gain in IPC confidence using logistic regression; the dependent variable was the outcome of whether the learner reported an increase in IPC confidence or not, and the independent variables of interest included course format, gender, profession, time since graduation, and number of completed course modules. A chi-squared test (two-sided) was used to compare study groups. Statistical significance was set at p<0.05. All analyses were conducted using Stata version 16.1.

### Ethics Statement

The design of the training program, including the topics covered and the format of the training, was informed by input from focus-group discussions with patient groups, learners (both pre-service and early career professionals), and HIV educators from a variety of settings in SSA, and has been previously described. [10, 14] Assessment tools to evaluate learners’ knowledge and confidence were also piloted with a subset of multidisciplinary learners before the full program was launched. All learners were given access to their pre- and post-score test results via the program’s learning management system (LMS). In addition, aggregate, site-level evaluation data were also posted on the LMS. The protocol for this project was reviewed and approved by the University of California, San Francisco’s Institutional Review Board (IRB) in San Francisco, California. Verbal consent was required at the time of participation in the study as approved by the IRB (protocol #: 19–28,447).

## Results

Between May 2021 and December 2021, 5,441 learners enrolled in the training program across 18 SSA countries; of whom complete demographic and evaluative data was available for 3842 learners that were included in the analysis. Nurses (n=1172, 37.9%) and physicians (n=825, 26.7%) comprised most of the participants (Table 1). The majority (67.2%) were pre-service learners (n=2072), and 13.0% (n=401) had graduated from training within the past 12 months; 176 learners participated in the in-person course, 2790 participated in the virtual workshop format, and 774 in the online course.

Across all modules, the mean 4-point Likert score in IPC confidence increased from 2.75 pre-participation to 3.29 post-participation (difference +0.54, p<0.001). Mean IPC confidence increased post-participation compared with pre-participation across each module with the smallest gain in the module ‘End-of-life Care for Patients with HIV’ (difference +0.28) and the largest gain from the module ‘Care for an Adolescent with Perinatally-acquired HIV’ (difference +1.10). Moreover, across all demographic variables of interest, there were increases in IPC confidence (Figure 2).

At the level of the individual learner, 68.3% (n=2622) reported gains in IPC confidence post-participation, while 17.7% (n=681) reported no change, and 14.0% (n=539) reported a reduction in their self-reported confidence. In unadjusted logistic regression, female gender (odds ratio [OR] 1.19, 95% Confidence Interval [CI] 1.03-1.37) was associated with greater odds of IPC confidence gain than male gender. Relative to nursing and midwifery professions, medical professionals (OR 2.12, 95% 1.73-2.60), and pharmacy professionals (OR 1.37, 95% CI 1.10-1.70) experienced greater odds of IPC confidence gain (Table 2). Moreover, completing more than four modules in a training course was associated with greater odds of IPC confidence gain (OR 2.25, 95% CI 1.96-2.58) than completing fewer modules. Participating in in-person workshops (OR 1.56, 95% CI 1.08-2.27) was also associated with significantly greater odds of IPC confidence gain compared to participation in blended online course format.

In multivariate logistic regression, adjusted for gender, training level, country, number of course modules completed and course format, female gender (OR 1.34, 95% CI 1.13-1.59), medical professional cadre (OR 1.96, 95% CI 1.56-2.46), having graduated greater than 12 months prior to participation (OR 1.46, 95% CI 1.49-4.39), and participating in virtual (OR 1.4, 95% CI 1.12-1.76) or in-person workshops (OR 2.55, 95% CI 1.49-4.39), were all associated with significant odds of IPC confidence gain (Table 2). Learners who completed four or more course modules also experienced significant odds of IPC confidence gain (OR 2.37, 95% CI 1.99-2.83).

## Discussion

Across 3842 interprofessional learners from 18 SSA countries, participating in a deliberate IPECP training program was associated with substantial gains in IPC confidence across gender, training level, health profession, country, number of course modules completed, and course format. In multivariate analysis, the odds of experiencing a gain in IPC confidence were greatest for women, medical professionals, those who participated in synchronous training programs (both in-person and virtual workshops), those who had been in professional practice for more than 12 months, and those who completed more than 4 modules. Moreover, this is also the largest interprofessional, multi-country training program aimed at improving HIV care quality among healthcare professionals in the region.[7] As such, these results have several policy and programmatic implications for the broader context of health professions education in Africa and for the provision of quality HIV care in SSA.

Firstly, this study underscores the importance of interprofessional education and collaborative practice (IPECP) in securing improved HIV care quality in programs such as the US President’s Emergency Plan for AIDS Relief (PEPFAR) in Africa. Africa has adopted the “treat all” approach [15] to HIV care and management which requires the efforts of nurses and midwives as part of a wider interprofessional team. Programs such as STRIPE HIV that not only equip learners with *knowledge* but also enhance IPC *confidence* are critical in the achievement of both HIV-related and broader healthcare goals and targets.[16] While acknowledging that gains in IPC confidence may not equate to greater teamwork or improved clinical outcomes, these data provide compelling evidence that team-based approaches to learning in SSA can improve provider confidence, which is a critical determinant of professional practice and clinical decision-making.[1-3, 5-7, 16-19] In addition to enhancing IPC confidence, we speculate that this intervention may have enhanced institutional capacity for IPECP at participating academic institutions across SSA. As such, future analyses will seek to determine whether the STRIPE HIV program increased educators’ confidence to deploy IPE approaches in other courses.

Secondly, the findings highlight the potential value of blended synchronous and asynchronous online and face-to-face educational models to enhance IPC confidence among healthcare professionals in SSA. While synchronous formats, both virtual and in-person increased IPC confidence, in-person training was associated with significantly greater gains in IPC confidence. Accordingly, these results validate findings from other recent training interventions that have highlighted both strengths and limitations of online learning modalities for health professions education in SSA.[20-22] In addition, the findings highlight the potential trade-offs between virtual synchronous and online, blended learning strategies, both associated with smaller IPC gains in our analysis, but likely to be less disruptive to clinical care, less expensive than in-person learning strategies, and more accessible to rural/remote providers. [23-25] More research is warranted to determine how to leverage digital tools to advance IPE in SSA. Although many health professional training institutions in Africa lack access and capacity to use digital technologies to deliver HIV training, [26] our results affirm the critical role that online, synchronous training can play in advancing IPC.

Thirdly we assert that the STRIPE HIV program offers a model for how to effectively enhance IPC confidence that can be leveraged to prepare for and respond to other public health and clinical challenges in SSA.[27] Moreover, our study endorses WHO recommendations in support of strengthening IPECP initiatives as critical to achieving universal health coverage [28, 29] and delivering team-based care beyond HIV care.[16, 30] As such, future research is critical to determine whether the learning modalities employed in this study can be employed to enhance collaboration between different professions in the delivery of other clinical programs and in response to other public health threats, such as future pandemics.[3, 31]

### Limitations

It is important to acknowledge limitations in the study, including self-reported confidence subject to social desirability bias, generalizability, and selection bias due to the inclusion of only those individuals who completed both pre-and post-tests, and only those who had access to the training in their setting. In addition, the study did not clearly establish whether the format of the course delivery (online vs. virtual workshop vs. blended online course) impacted learner engagement, an important consideration for future education and training initiatives in Africa. Finally, the findings do not provide evidence of improved teamwork or clinical outcomes, as these were not the focus of this evaluation. Nonetheless, they suggest that additional research, including examining the impact of IPE programs on clinical outcomes and cost-effectiveness, will be important. Evaluating the impact of interprofessional training programs, such as the STRIPE HIV program, to sustain and retain healthcare workers in clinical practice should also be a future priority.

## Conclusion

This is one of the largest interprofessional, multi-country training programs to improve HIV care among healthcare professional workers in SSA. These findings highlight the importance of in-person and virtual educational platforms to enhance IPC confidence. This analysis should inform curriculum development to strengthen care delivery across SSA.

## Author Contributions

All authors have approved the final version submitted.

## Data Availability Statement

Data included in this study are available in a public, open-access format on the Dryad dataset repository website: https://doi.org/10.7272/Q6WQ021N

---

**Figure 1.**
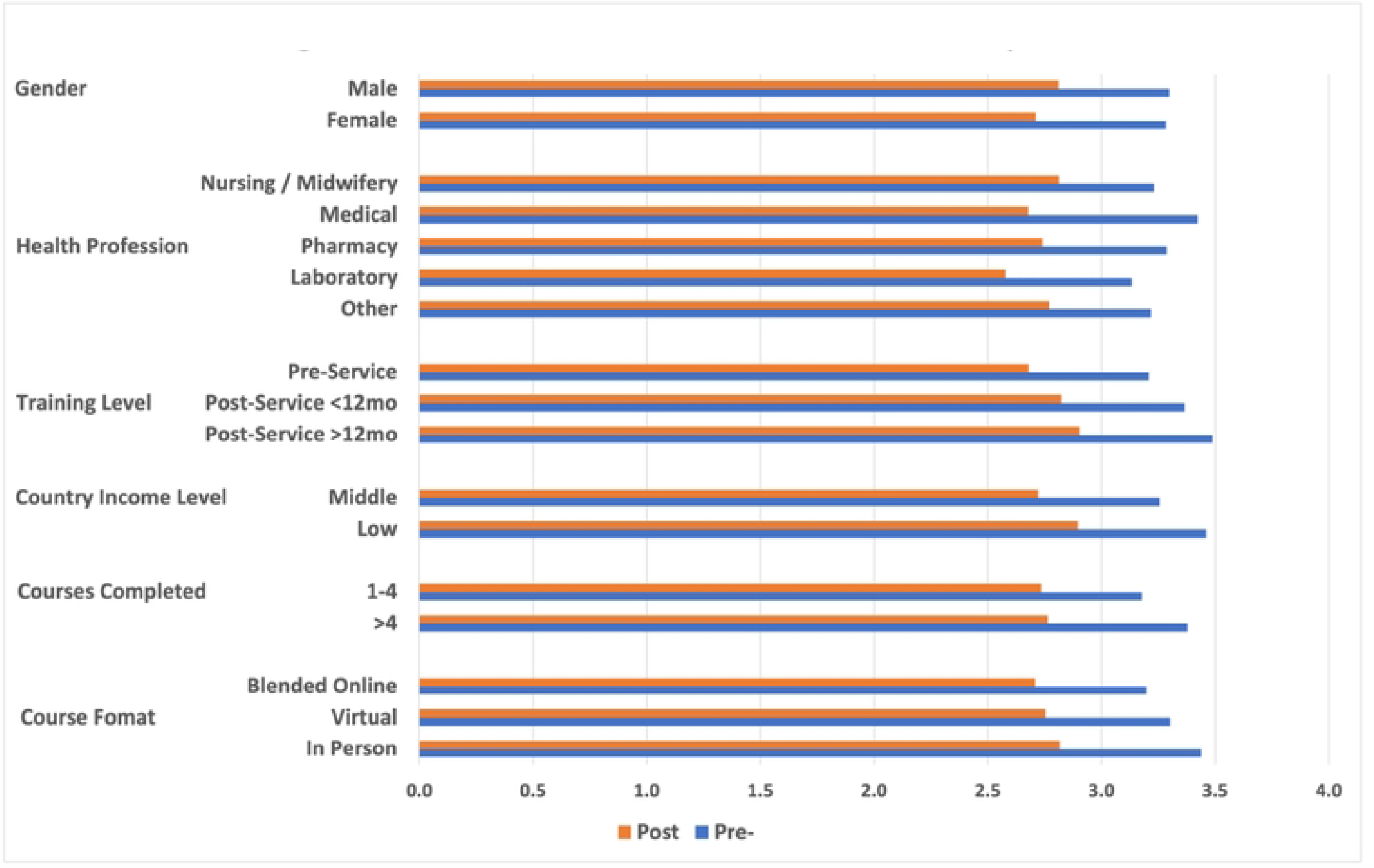
Mean IPC Confidence Score Pre- and Post-Participation.

## References

1. Reeves S, Zwarenstein M, Goldman J, Barr H, Freeth D, Hammick M, et al. Interprofessional education: effects on professional practice and health care outcomes. Cochrane Database Syst Rev. 2008;(1):CD002213. Epub 20080123. doi: 10.1002/14651858.CD002213.pub2. PubMed PMID: 18254002.

2. Reeves S. A systematic review of the effects of interprofessional education on staff involved in the care of adults with mental health problems. J Psychiatr Ment Health Nurs. 2001;8(6):533–42. doi: 10.1046/j.1351-0126.2001.00420.x. PubMed PMID: 11842481.

3. Hammick M, Freeth D, Koppel I, Reeves S, Barr H. A best evidence systematic review of interprofessional education: BEME Guide no. 9. Med Teach. 2007;29(8):735–51. doi: 10.1080/01421590701682576. PubMed PMID: 18236271.

4. Zwarenstein M, Bryant W, Bailie R, Sibthorpe B. [Meta-analysis of the Cochrane Collaboration. Promoting collaboration between nurses and physicians]. Assist Inferm Ric. 2000;19(2):97–9. PubMed PMID: 11107363.

5. Lemieux-Charles L, Chambers LW, Cockerill R, Jaglal S, Brazil K, Cohen C, et al. Evaluating the effectiveness of community-based dementia care networks: the Dementia Care Networks’ Study. Gerontologist. 2005;45(4):456–64. doi: 10.1093/geront/45.4.456. PubMed PMID: 16051908.

6. Mickan SM. Evaluating the effectiveness of health care teams. Aust Health Rev. 2005;29(2):211–7. doi: 10.1071/ah050211. PubMed PMID: 15865572.

7. Kitema GF, Laidlaw A, O’Carroll V, Sagahutu JB, Blaikie A. The status and outcomes of interprofessional health education in sub-Saharan Africa: A systematic review. J Interprof Care. 2023:1–23. Epub 20230205. doi: 10.1080/13561820.2023.2168631. PubMed PMID: 36739570.

8. Ahmat A, Okoroafor SC, Kazanga I, Asamani JA, Millogo JJS, Illou MMA, et al. The health workforce status in the WHO African Region: findings of a cross-sectional study. BMJ Glob Health. 2022;7(Suppl 1). doi: 10.1136/bmjgh-2021-008317. PubMed PMID: 35675966; PubMed Central PMCID: PMCPMC9109011.

9. Kiguli-Malwadde E, Budak JZ, Chilemba E, Semitala F, Von Zinkernagel D, Mosepele M, et al. Developing an interprofessional transition course to improve team-based HIV care for sub-Saharan Africa. BMC Med Educ. 2020;20(1):499. Epub 2020/12/11. doi: 10.1186/s12909-020-02420-x. PubMed PMID: 33298029; PubMed Central PMCID: PMCPMC7725217.

10. Kiguli-Malwadde E, Forster M, Martin S, Chilemba E, Couper I, Motlhatlhedi K, et al. Evaluating the impact of a multicountry interprofessional training programme to improve HIV knowledge and clinical confidence among healthcare workers in sub-Saharan Africa: a cohort study. BMJ Open. 2022;12(7):e060079. doi: 10.1136/bmjopen-2021-060079.

11. Omaswa F, Kiguli-Malwadde E, Donkor P, Hakim J, Derbew M, Baird S, et al. The Medical Education Partnership Initiative (MEPI): Innovations and Lessons for Health Professions Training and Research in Africa. Ann Glob Health. 2018;84(1):160–9. Epub 2019/03/16. doi: 10.29024/aogh.8. PubMed PMID: 30873813; PubMed Central PMCID: PMCPMC6586907.

12. Van Schalkwyk SC, Kiguli-Malwadde E, Budak JZ, Reid MJA, de Villiers MR. Identifying research priorities for health professions education research in sub-Saharan Africa using a modified Delphi method. BMC Med Educ. 2020;20(1):443. Epub 2020/11/20. doi: 10.1186/s12909-020-02367-z. PubMed PMID: 33208149; PubMed Central PMCID: PMCPMC7672834.

13. Omaswa F, Kiguli-Malwadde E, Donkor P, Hakim J, Derbew M, Baird S, et al. Medical Education Partnership Initiative gives birth to AFREhealth. Lancet Glob Health. 2017;5(10):e965–e6. Epub 2017/09/16. doi: 10.1016/S2214-109X(17)30329-7. PubMed PMID: 28911758; PubMed Central PMCID: PMCPMC5712669.

14. Kiguli-Malwadde E, Budak JZ, Chilemba E, Semitala F, Von Zinkernagel D, Mosepele M, et al. Developing an interprofessional transition course to improve team-based HIV care for sub-Saharan Africa. BMC Medical Education. 2020;20(1):499. doi: 10.1186/s12909-020-02420-x.

15. Nash D, Yotebieng M, Sohn AH. Treating all people living with HIV in sub-Saharan Africa: a new era calling for new approaches. J Virus Erad. 2018;4(Suppl 2):1–4. Epub 20181115. PubMed PMID: 30515307; PubMed Central PMCID: PMCPMC6248848.

16. WHO. Framework for Action on Interprofessional Education & Collaborative Practice. Geneva, Switzerland: WHO, 2010 2010. Report No.

17. Desender K, Boldt A, Yeung N. Subjective Confidence Predicts Information Seeking in Decision Making. Psychol Sci. 2018;29(5):761–78. Epub 20180402. doi: 10.1177/0956797617744771. PubMed PMID: 29608411.

18. Bandura A, Adams NE. Analysis of self-efficacy theory of behavioral change. Cognitive Therapy and Research. 1977;1:287–310.

19. Reeves S, Fletcher S, Barr H, Birch I, Boet S, Davies N, et al. A BEME systematic review of the effects of interprofessional education: BEME Guide No. 39. Med Teach. 2016;38(7):656–68. Epub 20160505. doi: 10.3109/0142159X.2016.1173663. PubMed PMID: 27146438.

20. Kagawa MN, Chipamaunga S, Prozesky D, Kafumukache E, Gwini R, Kandawasvika G, et al. Assessment of Preparedness for Remote Teaching and Learning to Transform Health Professions Education in Sub-Saharan Africa in Response to the COVID-19 Pandemic: Protocol for a Mixed Methods Study With a Case Study Approach. JMIR Res Protoc. 2021;10(7):e28905. Epub 20210728. doi: 10.2196/28905. PubMed PMID: 34254943; PubMed Central PMCID: PMCPMC8320735.

21. Enyama D, Balti EV, Simeni Njonnou SR, Ngongang Ouankou C, Kemta Lekpa F, Noukeu Njinkui D, et al. Use of WhatsApp(R), for distance teaching during COVID-19 pandemic: Experience and perception from a sub-Saharan African setting. BMC Med Educ. 2021;21(1):517. Epub 20211002. doi: 10.1186/s12909-021-02953-9. PubMed PMID: 34598681; PubMed Central PMCID: PMCPMC8486629.

22. Downie A, Mashanya T, Chipwaza B, Griffiths F, Harris B, Kalolo A, et al. Remote Consulting in Primary Health Care in Low- and Middle-Income Countries: Feasibility Study of an Online Training Program to Support Care Delivery During the COVID-19 Pandemic. JMIR Form Res. 2022;6(6):e32964. Epub 20220614. doi: 10.2196/32964. PubMed PMID: 35507772; PubMed Central PMCID: PMCPMC9200055.

23. Rowe SY, Peters DH, Holloway KA, Chalker J, Ross-Degnan D, Rowe AK. A systematic review of the effectiveness of strategies to improve health care provider performance in low- and middle-income countries: Methods and descriptive results. PLoS One. 2019;14(5):e0217617. Epub 20190531. doi: 10.1371/journal.pone.0217617. PubMed PMID: 31150458; PubMed Central PMCID: PMCPMC6544255.

24. Leslie HH, Gage A, Nsona H, Hirschhorn LR, Kruk ME. Training And Supervision Did Not Meaningfully Improve Quality Of Care For Pregnant Women Or Sick Children In Sub-Saharan Africa. Health Aff (Millwood). 2016;35(9):1716–24. doi: 10.1377/hlthaff.2016.0261. PubMed PMID: 27605655.

25. Ridde V. Per diems undermine health interventions, systems and research in Africa: burying our heads in the sand. Trop Med Int Health. 2010;15(7):E1–E4. doi: 10.1111/tmi.2607. PubMed PMID: 28639744.

26. Sharp A, Jain V, Alimi Y, Bausch DG. Policy and planning for large epidemics and pandemics - challenges and lessons learned from COVID-19. Curr Opin Infect Dis. 2021;34(5):393–400. doi: 10.1097/QCO.0000000000000778. PubMed PMID: 34342301; PubMed Central PMCID: PMCPMC8452318.

27. Reid M, Suleman F, De Villiers M. The SARS-CoV-2 pandemic: An urgent need to relook at the training of the African health workforce. S Afr Med J. 2020;110(4):12875. Epub 20200317. doi: 10.7196/SAMJ.2020.v110i4.14713. PubMed PMID: 32657733.

28. Agyepong I, Spicer N, Ooms G, Jahn A, Barnighausen T, Beiersmann C, et al. Lancet Commission on synergies between universal health coverage, health security, and health promotion. Lancet. 2023;401(10392):1964–2012. Epub 20230521. doi: 10.1016/S0140-6736(22)01930-4. PubMed PMID: 37224836.

29. Agyepong IA, Sewankambo N, Binagwaho A, Coll-Seck AM, Corrah T, Ezeh A, et al. The path to longer and healthier lives for all Africans by 2030: the Lancet Commission on the future of health in sub-Saharan Africa. Lancet. 2017;390(10114):2803–59. Epub 2017/09/18. doi: 10.1016/S0140-6736(17)31509-X. PubMed PMID: 28917958.

30. Organization WH. Transforming health workforce education in support of universal health coverage. Geneva: World Health Assembly, 2013 27 May 2013. Report No.

31. Thistlethwaite J. Interprofessional education: a review of context, learning and the research agenda. Med Educ. 2012;46(1):58–70. doi: 10.1111/j.1365-2923.2011.04143.x. PubMed PMID: 22150197.

